# Layer-specific cortical signatures uncover a sensory origin of post-stroke motor dysfunction

**DOI:** 10.64898/2026.07.30.26359200

**Authors:** Nan Liu, Lea Theresa Mais, Maike Mustin, Seong Dae Yun, Nadim Joni Shah, Gereon R. Fink, Christian Grefkes, Caroline Tscherpel

**Author notes:** Correspondence to*: Caroline Tscherpel, MD, PhD, Department of Neurology, University Hospital Frankfurt, Theodor-Stern-Kai 7, Frankfurt am Main, 60590, Germany.

## Abstract

The cerebral cortex computes through a laminar microcircuit in which superficial layers integrate cortico-cortical input and deep layers issue corticospinal output. How focal injury disrupts this input–output architecture has been characterized in animal models, but has never been resolved in the human brain, leaving it unknown whether stroke degrades the motor cortex uniformly or dissociates its computational compartments. We used ultra-high-field 7T functional MRI to resolve activation across cortical depth in the primary motor cortex (M1) hand knob during finger tapping (12 stroke patients/14 controls), and related layer-specific signals to comprehensive motor assessments through principal component analysis, correlation, and cross-validated predictive modeling. In the ipsilesional hemisphere, both the input (L2/3) and output (L5) layers showed reduced activation during affected-hand tapping movement in patients compared to controls, indicating a combined failure of sensorimotor integration and corticospinal output. In the contralesional hemisphere, the two compartments dissociated and tracked distinct behavioral processes: superficial (L1) activation varied with global motor outcome, whereas deep output-layer (L5/L6) activation tracked specifically with affected-hand grip force. Cross-validated modeling confirmed this double dissociation: superficial activation predicted global outcome, deep activation predicted grip force, and no ipsilesional layer predicted either. These findings provide the first human evidence that stroke does not disrupt the motor cortex uniformly but reorganizes at the level of individual laminar compartments, with the superficial input stage and deep output stage indexing separable aspects of motor dysfunction. Beyond linking cortical microcircuit models derived from animals to the organization of impairment in humans, layer-resolved imaging reveals prognostic information inaccessible to whole-region measures, pointing toward laminar signatures that could help stratify motor deficits and guide targeted rehabilitation.

## Introduction

The neocortex is organised into layers, and this layering is not incidental: it reflects a canonical microcircuit, conserved across areas and species, in which distinct layers receive, transform, and relay information along a stereotyped pathway (Douglas & Martin, 2004, 2007). In the primary motor cortex (M1), superficial and deep layers occupy opposite ends of this input– output axis (Harris & Shepherd, 2015), a functional segregation that underlies both healthy function and the response to injury. This laminar view is central to how motor cortex is understood in animals (Hooks et al., 2011; Weiler et al., 2008), but it has scarcely featured in human stroke research, which has largely remained at the scale of regions and networks.

This macroscale focus has shaped both what has been asked and what has been found. Previous structural and functional imaging studies have investigated the structural integrity, task-evoked responses, and functional connectivity of the motor network after stroke (Byblow et al., 2015; Grefkes & Fink, 2011, 2014; Rehme & Grefkes, 2013; Schulz et al., 2014, 2017; Wiemer et al., 2025). While ipsilesional cortical reorganisation has traditionally been considered critical for successful recovery post-stroke, the precise relevance of the contralesional motor cortex for reorganisation after stroke remains less conclusive. Some studies have demonstrated a less favorable motor outcome associated with increased contralesional hemisphere recruitment, whereas other evidence has suggested that contralesional activity may play an essential role in post-stroke motor recovery, determined by variables including time since stroke and complexity of motor task (Dodd et al., 2017; Matsuura et al., 2017). A possible source of this inconsistency is spatial scale. Conventional imaging measures the summed activity of a cortical column, collapsing functionally distinct layers, including the input and output compartments, into a single signal. If stroke engages these compartments differently, then the same increase in contralesional activation could reflect afferent input, or corticospinal output, each with different implications for recovery, which is an ambiguity that macroscale methods cannot resolve. Therefore, resolving cortical activation by layer would allow one to ask not merely whether a region is recruited after stroke, but which laminar processes that recruitment reflects, and which patterns of reorganisation are more likely to support motor recovery.

For the well-characterised somatotopic organisation of the motor system, axonal tracing studies have demonstrated that the six layers of the neocortex are interconnected through canonical microcircuits that support information processing across cortical depths (Douglas & Martin, 2007; Norris & Polimeni, 2019). This inter-laminar connectivity of M1 can be conceptualized as a two-loop system (Weiler et al., 2008). Within this framework, the upper loop, encompassing layers I–III, is primarily driven by external excitatory input through cortico-cortical and thalamo-cortical projections (Ninomiya et al., 2019; Papale & Hooks, 2018). Subsequently, signals are transmitted via pyramidal neurons projecting from layer III to layer V, thereby engaging a lower loop comprising layers V–VI, which is predominantly responsible for generating corticospinal output (Kaneko et al., 2000).

Whether this laminar pattern is mirrored in the human brain has been beyond reach until recently. Recent advances in functional ultra-high-field magnetic resonance imaging (UHF-MRI) have opened new avenues for measurement techniques and analytical approaches, thereby enhancing our understanding of human brain function. Owing to its increased signal-to-noise ratio, 7T functional magnetic resonance imaging (fMRI) enables mesoscopic imaging at sub-millimeter spatial resolution, providing insights that were previously attainable only through ex-vivo studies or in animal models (De Martino et al., 2018). This enhanced resolution not only facilitates a more refined topological delineation of neuronal structures but also allows neuronal activity to be mapped across different cortical depths, offering a deeper understanding of the brain’s functional organisation (Huber et al., 2020). This makes it possible to differentiate between input-related activity in the superficial (supragranular) layers and output-related activity in the deeper (infragranular) layers of the cortex (Persichetti et al., 2020; Polimeni & Uludağ, 2018), linking cortical circuit principles established in animals to human cognition (Lawrence et al., 2019; McColgan et al., 2020a; Yang et al., 2021). Huber et al. (2017) demonstrated it directly in human M1, resolving the input-output compartments in vivo during finger-tapping movements. Consistent with this distinction, hand movements evoke responses in both superficial and deep layers, whereas touch sensation, which lacks motor output, involves only superficial layers (Persichetti et al., 2020).

This layer-specific functional organisation raises an important question that has not yet been addressed: does damage to motor circuits disrupt specific cortical layers distinctively? Animal studies reveal that post-stroke reorganisation is not uniform across cortical depth but instead engages the input and output loops in distinct ways. Within the upper loop, peri-infarct layer 2/3 pyramidal neurons show rapid dendritic spine loss within hours of ischemic injury, with surviving spines becoming elongated (Brown et al., 2008), alongside altered excitability. Tonic GABAergic inhibition rises in peri-infarct layer 2/3 pyramidal neurons in the weeks after stroke, and reducing it enhances motor recovery, marking supragranular excitability as a key constraint on reorganisation (Clarkson et al., 2010). The lower loop is affected differently: layer 5 pyramidal neurons in peri-infarct cortex show a comparable early loss of dendritic spines (Brown et al., 2008), which recovers gradually over subsequent weeks in step with local reperfusion, reflecting fine-scale synaptic plasticity (Mostany et al., 2010). However, their dendritic arbors show no evidence of compensatory growth, instead undergoing a two-step pruning process: initial retraction followed by branch loss (Mostany & Portera-Cailliau, 2011). This deep-layer remodeling may also be bilateral: contralesional layer 5 pyramidal neurons have been reported to grow additional apical dendrites after unilateral injury, though this finding remains contested and has no reliably established link to behavioral outcome (Jones & Adkins, 2015). Similar layer-specific alterations in M1 activation have been reported in other motor diseases: a recent laminar study of patients with focal hand dystonia revealed increased activity in input layers (L2/3) and reduced activity in the deep output layers (L5) of M1 during finger tapping, suggesting that motor disorders can produce distinct layer-specific dysfunction patterns (Huber et al., 2023).

While UHF-MRI has been used for structural, vascular, and microvascular imaging, for example, depicting ischemic lesions, vessel-wall pathology, and microbleeds at high spatial resolution (De Cocker et al., 2018; Kurz et al., 2025), layer-dependent functional imaging of reorganisation after stroke and post-stroke cortical dysfunction has not been achieved. To this end, we leveraged laminar fMRI at 7T to provide the first characterization of layer-specific activation patterns in the M1 hand knob region during finger tapping in chronic stroke patients. In combination with behavioral assessment, we aimed to identify whether recovery-related activity is primarily driven by increased afferent input in superficial layers or by enhanced output generation in deep layers. In detail, we examined (i) whether stroke-related activation changes in the ipsilesional M1 are uniform across cortical layers or whether superficial and deep layers show differential alterations after stroke, (ii) whether contralesional overactivity known from 3T fMRI studies could be replicated with 7T and, if so, whether it shows layer-specific patterns, and (iii) how layer-specific activation patterns in the motor cortex link with motor control and outcome after stroke.

## Methods and Materials

### Participants

Twelve stroke patients with unilateral upper limb motor deficits ranging from mild to severe (1 female, 11 males; 11 right-handed; mean age 61 ± 11 years (mean ± standard deviation, SD), range 40-81) were included in this study from the Department of Neurology, University Hospital of Cologne. Detailed participant demographics and clinical characteristics are presented in Table 1. Fourteen age-matched healthy participants without any history of neurological or psychiatric disorders (4 females, 10 males; 13 right-handed; mean age 63 ± 13 years, range 38-78) were included as a control group. Three patients and one control participant were initially scanned but were excluded from this study due to head movement exceeding ±2mm during the task. All participants provided written informed consent prior to participation. The study was approved by the ethics committee of the medical faculty at the University of Cologne (file no. 21-1419) and conducted in accordance with the Declaration of Helsinki.

**Table 1.**
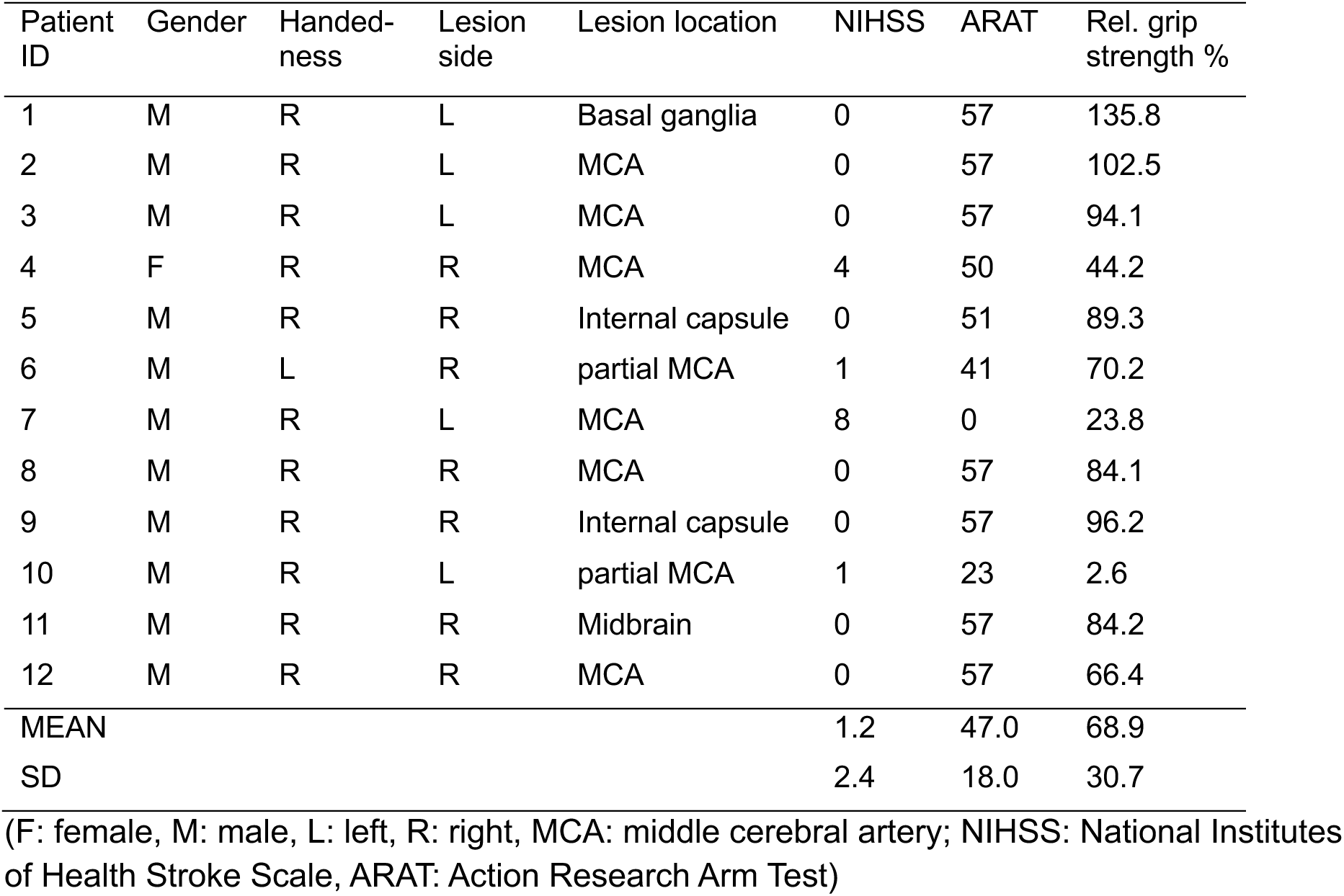
Demographic and clinical parameters of stroke patients.

### Experimental design

We employed an fMRI block design with a simple finger tapping task. Participants were instructed to perform repetitive finger tapping movements as fast as possible with their left or right index finger upon presentation of a visual cue. The visual cues, which were arrows pointing to either left or right, were presented in the center of the screen using the Presentation software (Neurobehavioral Systems, https://www.neurobs.com/) and instructed participants to perform the corresponding finger tapping movement. The experiment consisted of 14 blocks, each beginning with a 30-second baseline period, followed by an alternating sequence of five 3.8-second tapping trials and four 2.8-second pauses (total block duration: approximately 60 seconds). The index finger used for tapping (left or right) alternated across blocks, and block order was randomized across participants. The complete task lasted approximately 15 minutes.

### Behavioral assessment

We assessed six reliable motor and clinical parameters: (i) Maximum grip force was measured separately for each hand in three consecutive trials using a vigorimeter (KLS Martin Group, Germany). For each participant, the absolute grip force of each hand was calculated as the mean of three trials. Relative grip strength was additionally calculated and used for further analysis, defined as the ratio of the stroke-affected hand’s mean grip force (or the dominant hand for controls) to the unaffected hand’s mean grip force (or the non-dominant hand for controls) multiplied by 100; (ii) Purdue Pegboard Test evaluated arm and hand function and finger dexterity (Reddon et al., 1988). Participants were instructed to insert as many pins as possible into designated holes on a board within 30 s for three trials with each hand. The average of three trials per hand was computed for further analysis; (iii) Jebsen-Taylor hand function test (JTT) evaluated upper limb function through simulated activities of daily living (Jebsen et al., 1969). Participants completed six standardized tasks as quickly as possible, including turning over cards, picking up small objects, simulated feeding, stacking checkers, lifting large lightweight objects, and lifting large heavy objects. The total completion time for all tasks was calculated for each hand and inverted to maintain consistency with other motor test scores (higher scores = better performance); (iv) Action research arm test (ARAT) evaluated gross and fine upper limb function and related disabilities in neurological patients (Hsieh et al., 1998; Lyle, 1981). It consists of grasping, gripping, pinching, and gross movement tasks, accounting for different aspects of upper limb motor function; (v) Fugl-Meyer Assessment (FMA) scale, a widely used clinical evaluation tool designed to assess upper and lower extremity motor functions and recovery in stroke patients (Fugl-Meyer et al., 1975); (vi) National Institutes of Health Stroke Scale (NIHSS) assessed global neurological impairment. Based on these parameters, we also calculated a motor composite score for further analysis (see *Principal Component Analysis section for details*).

### MRI data acquisition

High-resolution images were acquired using a 7T Siemens Magnetom Terra MRI scanner (Siemens Healthineers, Erlangen, Germany), equipped with a 32-channel head coil. T1-weighted anatomical images were obtained using a three-dimensional magnetization-prepared rapid gradient-echo (MP2RAGE) sequence (Parameters: repetition time (TR) = 4300 ms, echo time (TE) = 2.08 ms, two inversion times (TI1 = 840 ms, TI2 = 2370 ms), flip angles of 5° and 6° for the first and second inversion, respectively, field of view = 240 × 225 mm, matrix size = 400 × 376, voxel size = 0.6 × 0.6 × 0.6 mm³ isotropic). The MP2RAGE sequence generated both individual inversion images (INV1 and INV2) and a unified T1-weighted image (UNI) with optimized contrast and reduced B1 inhomogeneity artifacts. The total acquisition time for the anatomical scan was approximately 8 minutes.

Functional MRI images were acquired using repetition-time-external (TR-external) echo-planar imaging with keyhole (EPIK) method (Yun et al., 2022, 2024). The parameters are as followings: repetition time (TR) = 3500 ms, echo time (TE) = 22 ms, flip angle = 85°, field of view = 212 × 212 mm^2^, matrix size = 336 × 336, slice thickness = 0.63 mm with no gap, voxel size = 0.63 × 0.63 × 0.63 mm³ isotropic. A total of 132 axial slices were acquired with a -9.2° tilt from the anterior-posterior commissure line to reduce susceptibility artifacts in orbitofrontal regions. The sequence employed 3-fold in-plane (GRAPPA) and 3-fold inter-plane (multi-band) acceleration to achieve high spatial and temporal resolution. Both magnitude and phase images were collected simultaneously for potential distortion correction and improved signal processing. Partial Fourier encoding (6/8) was used to reduce acquisition time while maintaining image quality. Pulse and breathing were monitored during the acquisition using a finger oximeter and a respiratory belt. The acquisition time of the whole functional scan was approximately 15 minutes.

### MRI data analysis

#### Preprocessing

T1-weighted anatomical images underwent a multi-step preprocessing pipeline to optimize image quality and enable accurate tissue segmentation. First, denoising was performed using LayNii (Huber et al., 2021) to remove background noise commonly associated with MP2RAGE sequences at ultra-high field strengths. Subsequently, bias field correction was applied using SPM12 (Statistical Parametric Mapping, Wellcome Trust Centre for Neuroimaging, London, UK) through joint estimation of bias field inhomogeneities and tissue segmentation using a Gaussian mixture model approach. To prepare MP2RAGE data for further segmentation processing, PreSurfer pipeline (https://github.com/srikash/presurfer) was employed to enhance contrast properties and generate brain extraction masks from the INV2 image, which were subsequently applied to clean up the non-brain parts of the preprocessed UNI image. Whole-brain segmentation was performed using FreeSurfer (Fischl, 2012), with the preprocessed UNI image and brain-extracted masks as input for comprehensive cortical surface reconstruction and subcortical segmentation. This preprocessing pipeline was specifically tailored to address the unique challenges of 7T MP2RAGE data while maximizing the spatial resolution advantages of ultra-high field imaging (Pizzuti et al., 2025). FreeSurfer’s subcortical segmentation was then converted to cortical rim masks and upsampled to 0.2 mm isotropic resolution using nearest neighbor interpolation in Convert3D from ITK-SNAP (version 4.2.2, https://www.itksnap.org/pmwiki/pmwiki.php) for further layerification analysis.

Functional MRI data, including magnitude and phase images, were first slice-timing corrected to the middle slice using SPM12, followed by motion correction using AFNI (https://afni.nimh.nih.gov/) with a middle volume as reference. Processed phase images were unwrapped and used to generate fieldmaps, which were then applied to correct for susceptibility-induced geometric distortions using FSL (Jenkinson et al., 2012). Functional-to-anatomical registration was performed using FreeSurfer’s boundary-based registration (bbregister) optimized for BOLD contrast. Brain masks were generated from the processed functional data using AFNI (3dAutomask) to exclude non-brain voxels from subsequent analyses. Finally, general linear models (GLMs) were used to estimate BOLD responses (β values) to left and right finger tapping conditions, with polynomial detrending up to 7th order to remove low-frequency drift. Head motion parameters derived from the motion correction step were included as nuisance regressors to account for residual motion-related variance. Beta coefficient maps were subsequently extracted for left and right finger tapping conditions and converted to percent signal change maps. To match the resolution of the anatomical cortical rim masks, these maps were upsampled to 0.2 mm resolution using cubic interpolation and masked by the upsampled cortical rim masks to exclude white matter (Huber et al., 2021). After preprocessing, we assessed the quality of anatomical images for each participant and evaluated head-motion parameters individually. For patients with right-hemisphere lesions, MRI images were flipped left-to-right across the mid-sagittal plane after preprocessing, ensuring that the lesion was consistently located in the left (‘ipsilesional’) hemisphere for all subsequent analyses. Motor measures (e.g., ARAT, Fugl-Meyer, grip strength) were relabeled by laterality (left/right) to match this flip, ensuring that “affected hand” consistently corresponded to the hand contralateral to the (now left-sided) lesion across all patients.

#### Layerification

Cortical layerification was performed using LayNii (Huber et al., 2021). We used the upsampled cortical rim masks to create equidistant layers spanning from the white matter to the pial surface. To ensure complete coverage across the cortical ribbon, complementary leaky layers were computed to fill any gaps in the layer structure. The final cortical depth model combined both layer types with spatial smoothing (1 mm FWHM) to produce 20 discrete layers for subsequent layer-specific analysis (Huber et al., 2016; Huber et al., 2021). The 20 layers were used based on recommendations from recent 7T studies for optimal visualization of layer profiles in the motor cortex (Huber et al., 2017; Persichetti et al., 2020). Given the 0.2 mm resolution and estimated 4 mm cortical thickness in the motor cortex, this approach samples approximately 20 voxels across the cortical depth, with roughly 1 voxel per layer. Layer-specific smoothing was then applied to the functional activation maps (0.4 mm FWHM) using LayNii’s LN_LAYER_SMOOTH to preserve cortical depth information while reducing noise.

#### Region of interest (ROI) selection and columnar analysis

As the main focus of this study was about motor reorganisation of the upper limb, we focused on the layer-specific profiles in the hand knob area. Therefore, we first created bilateral M1 masks from the FreeSurfer DKT atlas parcellation (Klein & Tourville, 2012) and upsampled them to 0.2 mm resolution to match the resolution used for layerification. Mid-gray matter layers (layers 10-11) were isolated to define the cortical column reference. Cortical columns were then generated within M1 using LayNii’s LN2_COLUMNS, creating 200 vertical columns per hemisphere. This approach utilized 3D cortical geometry to enable sampling of layer-specific activation patterns within anatomically defined cortical columns in M1. We selected the hand knob ROIs within columnized M1 using two criteria. First, the generated cortical columns within M1 were reviewed by a senior neurologist to identify those located within the hand knob region based on its unique anatomical landmarks for each subject. Secondly, columns within these anatomically defined areas that overlapped with peak activation to contralateral finger tapping were selected as ROIs. This resulted in 3-4 columns per hemisphere as hand knob ROIs. These ROIs were labeled as lateral, middle, and medial hand knob subregions based on their anatomical location (see an illustration in Figure 1A). We focused primarily on the lateral cortical columns, as thumb and index finger representations are known to be laterally localized in the hand knob area (Beisteiner et al., 2001; Schweisfurth et al., 2018; Siero et al., 2014). Cortical layers within ROIs were further grouped based on cortical depth as follows: L1 (0–20% cortical depth), L2/3 (20–50%), L5 (50–67%), and L6 (67–100%) (Nothnagel et al., 2025). Layer-specific BOLD activation profiles were subsequently extracted from each cortical layer within lateral hand knob ROIs for the control and patient groups, respectively (see an illustration in Figure 1A).

**Figure 1.**
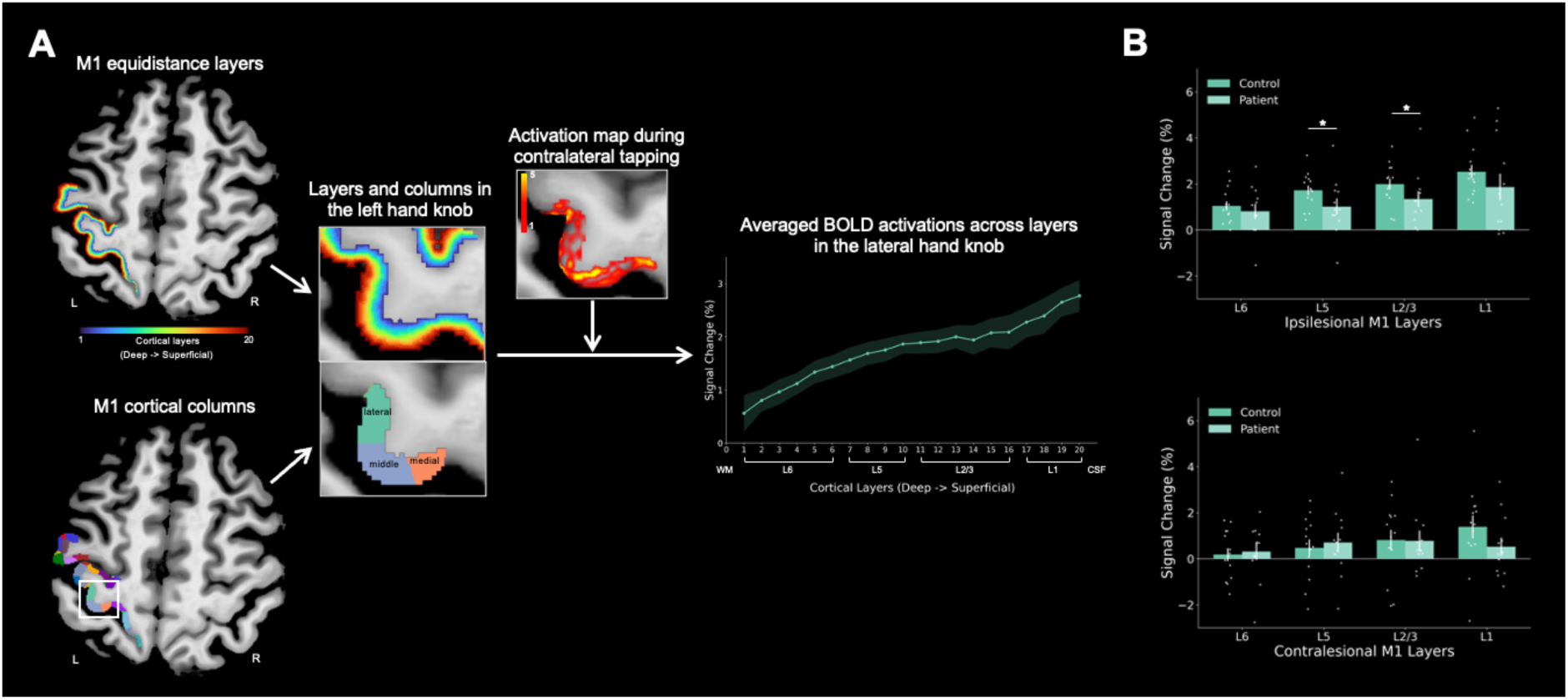
Layer-specific BOLD activation in the hand knob region during finger tapping in chronic stroke patients and healthy controls. **A**. Illustration of cortical layer segmentation and columnization in the left primary motor cortex (M1) from a representative healthy participant. Cortical layers were defined as: L1 (0–20% cortical depth), L2/3 (20–50%), L5 (50–67%), L6 (67–100%). The line plot shows layer-specific BOLD activation profiles in the left lateral hand knob during contralateral (right) finger tapping, averaged across all controls (n = 14). Shaded areas represent the standard error of the mean (SEM). **B**. Comparison of layer-specific BOLD activation of the ipsilesional (left) and contralesional (right) lateral hand knob during finger tapping of the affected (right) hand between controls and chronic stroke patients (14 controls/12 patients). Both L2/3 and L5 showed significantly reduced activation in the ipsilesional hemisphere in patients compared with controls, and no layer showed a significant group difference in the contralesional hemisphere. Asterisks (*) indicate significant between-group differences (* for *p* < 0.05).

#### Principal Component Analysis (PCA)

To identify the underlying components driving performance across motor tests and to extract a functional motor composite score, we performed PCA on z-transformed motor scores from all patients. The dataset included four motor assessment parameters: Purdue Pegboard Test scores, inverted JTT scores, ARAT scores, and Fugl-Meyer Assessment scores of the affected (right) hand. Grip force was not entered into the PCA because it reflects maximal voluntary strength, which differs from the dexterity- and function-based measures and forms a separable dimension in the data. Therefore, it was examined separately through the individual correlation analyses described below. We then calculated individual component scores for each patient using regression, indicating how well each patient performed on the motor abilities represented by each component.

#### Correlation Analysis

Following PCA, we computed Pearson correlations between the component scores of each extracted principal component (PC) and laminar-specific BOLD activation across layers (L1, L2/3, L5, L6) of the lateral hand knob during finger tapping of the affected hand, separately for the ipsilesional and contralesional hemispheres, in the patient group. To further assess clinical severity-related layer activation profiles in the lateral hand knob areas of stroke patients, we performed additional correlation analyses examining relationships between layer-specific BOLD activation of affected-hand finger tapping and a set of clinical measures, including ARAT and Fugl-Meyer Assessment scores of the affected hand, NIHSS scores, absolute grip strength of the affected hand, and relative grip strength. Pearson correlations were computed separately for each cortical layer in the ipsilesional and contralesional lateral hand knob areas, and FDR correction was applied for the four layers. Since absolute grip strength of the affected hand is correlated with relative grip strength, we complemented these zero-order correlations with partial correlations, in which each grip measure was related to layer-specific activation while controlling for the other, thereby isolating the variance uniquely attributable to each. Partial coefficients were derived from the three pairwise zero-order correlations using the standard first-order formula:

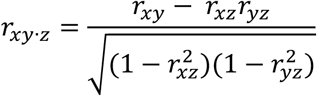

where *x* and *y* denote the grip measure and layer activation, and *z* the controlled grip measure. Comparing the zero-order and partial coefficients allowed us to determine whether a given grip–activation association was independent of, or instead driven by, the shared variance between the two grip measures.

#### Exploratory Predictive Modeling Analysis

To further explore whether cortical layer-specific activation profiles could predict motor outcomes in stroke patients, we conducted predictive modeling using a conservative leave-one-out cross-validation (LOOCV) framework given the limited sample size (12 patients). We focused on layer-specific activation profiles during finger tapping of the affected hand in the ipsilesional and contralesional lateral hand knob. For each combination of hemisphere and cortical layer (L1, L2/3, L5, L6), we constructed linear regression models with motor outcomes as dependent variables and layer-specific BOLD activation as predictors. Outcomes included PCA-derived component scores and individual clinical motor assessments (ARAT scores, Fugl-Meyer scores of the affected hand, NIHSS, and absolute grip strength of the affected hand).

#### Statistical Analysis

Statistical analyses were conducted using customized Python scripts. To determine which layer of the lateral hand knob was affected by stroke during finger tapping of the affected hand, we compared BOLD activations between controls and patients using two-tailed Wilcoxon rank-sum tests for each cortical layer (L1, L2/3, L5, L6) in the ipsilesional (left) and contralesional (right) hemispheres separately. Significance was assessed by permuting group labels (5000 iterations) to generate a null distribution of the rank-sum statistic, and the resulting p-values were FDR-corrected across the four layers within each hemisphere. Effects were considered significant at p_FDR < 0.05.

The Kaiser-Meyer-Olkin (KMO) measure of sampling adequacy and Bartlett’s test of sphericity were computed to verify that the dataset met the assumptions for PCA. Principal Components (PCs) were extracted using the Kaiser criterion (eigenvalues > 1), and a varimax orthogonal rotation was applied to maximize interpretability of the component structure (Schmidt et al., 2000; Wunderle et al., 2024). Component loadings above 0.4 were considered reliable, and loadings above 0.7 were regarded as meaningful for component interpretation.

For the correlation between layer-specific activation and both the PC scores and the individual clinical measures, Pearson correlation p-values were FDR-corrected for multiple comparisons across the four layers, separately for each component or measure within each hemisphere (two-tailed, p_FDR < 0.05). For the partial correlation, the significance of each partial coefficient was tested with:

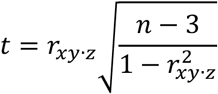

evaluated against Student’s *t* distribution with *n* − 3 degrees of freedom (two-tailed). The resulting p-values were then FDR-corrected across the four layers within each grip measure and hemisphere, with significance at p_FDR < 0.05.

In predictive modeling analyses, model performance was evaluated using LOOCV to provide unbiased estimates of predictive accuracy. Statistical significance of predictive models was also determined through permutation testing with 10,000 iterations, where outcome variables were randomly shuffled to generate null distributions of R² values, with p-values calculated as the proportion of permuted R² values greater than or equal to the observed R² value. We incorporated the standard add-one correction, i.e., ([number of permuted R² ≥ observed] + 1) / (number of permutations + 1), to ensure a valid non-zero p-value (Phipson & Smyth, 2010). For each model, we computed training R² and cross-validated R² to assess potential overfitting, with the overfitting gap (training R² minus cross-validated R²) serving as an indicator of model stability.

## Results

### Comparison of layer-specific profiles between control and patient groups in the lateral hand knob

To identify which cortical layers showed stroke-related alterations within the lateral hand knob, we compared BOLD activation during finger tapping of the affected (right) hand between controls and patients separately for each layer (L1, L2/3, L5, L6) and hemisphere (ipsilesional/left and contralesional/right) using two-tailed permutation-based Wilcoxon rank-sum tests. Stroke-related reductions were observed in the ipsilesional, not the contralesional, lateral hand knob (Fig. 1B). In the ipsilesional hemisphere, chronic stroke patients showed significant reduced BOLD activity in L2/3 and L5 (L2/3: U = 125, *p* = 0.018, Cohen’s d = 0.63; L5: U = 124, *p* = 0.021, Cohen’s d = 0.70; both *p_FDR* = 0.043; Fig. 1B, upper panel), whereas L1 and L6 did not differ significantly (L1: U = 111, *p* = 0.088, Cohen’s d = 0.42, *p_FDR* = 0.118; L6: U = 93, *p* = 0.330, Cohen’s d = 0.25; *p_FDR* = 0.330). No layer in the contralesional lateral hand knob showed a significant reduction (Fig. 1B, lower panel; see statistical details in Table S1).

### PCA and layer-specific activation profiles in the lateral hand knob

To examine the relationships between motor performance and layer-specific activation profiles in the lateral hand knob, we conducted PCA on motor performance measures and correlated the resulting components with layer-specific BOLD activations during finger tapping of the affected hand. PCA revealed two PCs that together explained 94.62% of the total variance. The first component PC1 - termed “motor outcome” - showed strong positive loadings from affected-hand ARAT and Fugl-Meyer (Fig. 2A, left column). This component represents overall motor outcome, and its scores in the patient group were significantly negatively correlated with superficial (L1) layer activation in the contralesional lateral hand knob (L1: r = -0.746, *p* = 0.005, *p_FDR* = 0.021) during finger tapping of the affected hand (Fig. 2B, upper row; Table S2). This indicated that higher activation of the superficial layer (L1) in the contralesional lateral hand knob was associated with greater motor deficits. The second component, PC2 - termed “fine motor” - reflected fine motor dexterity, with strong positive loadings from affected-hand Pegboard and inverted JTT scores (Fig. 2A, right column). It was not associated with layer activation in either the ipsilesional or the contralesional lateral hand knob (Fig. 2B, lower row; Table S2). Together, these results reveal that the stroke-related motor outcome component was associated with elevated superficial-layer (L1) activation in the contralesional lateral hand knob during affected-hand finger movement, with greater activation reflecting poorer motor function, whereas fine motor dexterity showed no relationship with activation in either hemisphere.

**Figure 2.**
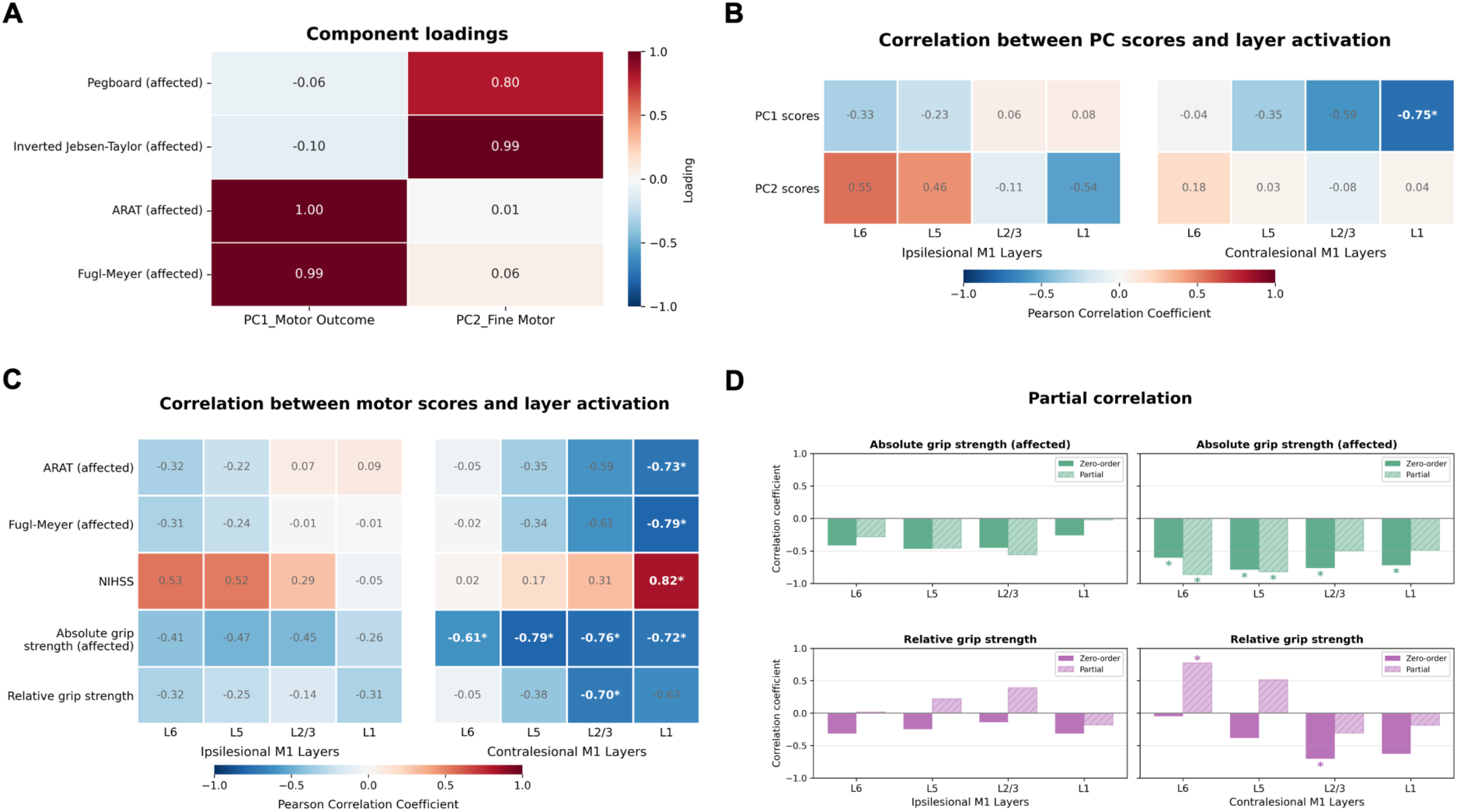
Relationships between layer-specific activation in the lateral hand knob and clinical motor outcomes after stroke. **A**. Principal component analysis (PCA) of motor assessments. The first two components, which together explained 94.62% of the variance, were retained. PC1 (“motor outcome”) showed strong positive loadings from ARAT and Fugl-Meyer scores of the affected (right) hand. PC2 (“fine motor”) showed strong positive loadings from Pegboard and inverted Jebsen-Taylor scores of the affected hand (Jebsen-Taylor time was inverted so that higher scores reflect better performance, consistent with the other measures). **B**. Pearson correlation between PC1/PC2 component scores and layer-specific BOLD activation during finger tapping of the affected hand in the ipsilesional and contralesional lateral hand knob in patients. Color indicates correlation direction and strength (blue: negative; red: positive). **C**. Pearson correlation between individual clinical measures and layer-specific BOLD activation during finger tapping of the affected hand in the ipsilesional and contralesional lateral hand knob in patients. The clinical measures include ARAT and Fugl-Meyer scores of the affected hand, NIHSS, absolute grip strength of the affected hand, and relative grip strength. **D**. Partial correlations between absolute and relative grip force and layer-specific activation, with each grip measure controlling for the other, given their intercorrelation. Asterisks represent significant correlations (\**p* < 0.05).

### Individual clinical measures and layer-specific activation profiles in the lateral hand knob

In case the PCA components did not fully capture stroke-related aspects, we directly correlated clinical assessments (ARAT and Fugl-Meyer scores of the affected hand, NIHSS, absolute grip strength of the affected hand, and relative grip strength) with layer-specific activations in the lateral hand knob during finger tapping of the affected hand in patients. ARAT scores of the affected hand correlated negatively with L1 activation of the contralesional lateral hand knob during finger tapping of the affected hand (r = -0.733, *p* = 0.007, *p_FDR* = 0.027), indicating that higher L1 activation was associated with worse functional motor performance (Fig. 2C). Fugl-Meyer scores showed the same negative correlation with L1 activation in the contralesional lateral hand knob during finger tapping of the affected hand (r = -0.790, *p* = 0.002, *p_FDR* = 0.009), with higher L1 activation related to lower Fugl-Meyer scores and worse motor performance (Fig. 2C). In line with that, NIHSS scores correlated positively with L1 activation in the contralesional lateral hand knob during finger tapping of the affected hand (r = 0.823, *p* = 0.001, *p_FDR* = 0.004), confirming that higher L1 activation was associated with greater stroke severity (Fig. 2C). These findings indicate that distinct clinical measures consistently revealed a pattern that higher contralesional superficial layer activation was related to worse clinical outcomes. Regarding grip strength, we found that activation of all the layers in the contralesional lateral hand knob during finger tapping with the affected hand was negatively correlated with the absolute grip strength of the affected hand (all *p_FDR* < 0.05, see statistical details in Table S3), whereas only L2/3 activation in the contralesional lateral hand knob showed significant negative correlation with relative grip strength (r = -0.704, *p* = 0.011, *p_FDR* = 0.043). This suggested that greater activation across all layers in the contralesional lateral hand knob was associated with weaker grip strength of the affected hand in chronic stroke patients, whereas relative grip strength showed a significant association with L2/3, with higher activation related to worse relative grip strength.

Given that absolute and relative grip strength are correlated, we complemented these correlations with partial correlations, in which each grip measure was related to layer-specific activation while controlling for the other. After controlling for relative grip strength, the negative correlation between contralesional activation and absolute grip strength of the affected hand remained significant and was slightly strengthened in the deep output layers L5 and L6 (L5: r = −0.82, *p* = 0.002, *p_FDR* = 0.004; L6: r = −0.87, *p* = 0.0006, *p_FDR* = 0.002), whereas L1 and L2/3 were no longer significant (L1: r = −0.49, *p* = 0.126; *p_FDR* = 0.126; L2/3: r = -0.50, *p* = 0.115; *p_FDR* = 0.126; Fig. 2D, upper right panel). Conversely, after controlling for absolute grip strength of the affected hand, the L2/3 association with relative grip strength observed in the simple correlation was abolished (r = −0.31, *p* = 0.358, *p_FDR* = 0.477), and the only significant partial correlation was a positive association at L6 (r = 0.78, *p* = 0.005, *p_FDR* = 0.020; Fig. 2D, lower right panel). These results indicate that the link between elevated contralesional activation and weaker grip was attributable specifically to absolute grip strength of the affected hand and localized to the deep output layers (L5/L6), independent of relative grip strength. The L2/3–relative-grip relationship, in contrast, was not independent of absolute strength, suggesting it reflected shared variance with absolute grip rather than a relative-strength–specific effect.

### Predictive modeling of motor outcomes from layer-specific activation

To assess the predictive capability of layer-specific activation patterns for motor outcome, we conducted LOOCV analyses in patients. Based on the correlation results above, we predicted PC1 motor outcome component and individual clinical measures (ARAT and Fugl-Meyer scores of the affected hand, NIHSS, and absolute grip strength of the affected hand) from layer-specific activation profiles in the patient group. The predictive results are shown in Figure 3. L1 activation in the contralesional lateral hand knob during finger tapping of the affected achieved significant cross-validated prediction performance for PC1 (“motor outcome”) (R² = 0.118, *p* = 0.030; Fig. 3A). Moreover, analysis of individual clinical measures confirmed the predictive capability of superficial layer (L1) activation in the contralesional lateral hand knob in patients. Specifically, L1 activation during finger tapping of the affected hand established significant predictive capacity for ARAT (R² = 0.086, *p* = 0.016; Fig. 3B), Fugl-Meyer (R² = 0.234, *p* = 0.006; Fig. 3C), and NIHSS scores (R² = 0.318, *p* = 0.001; Fig. 3D).

**Figure 3.**
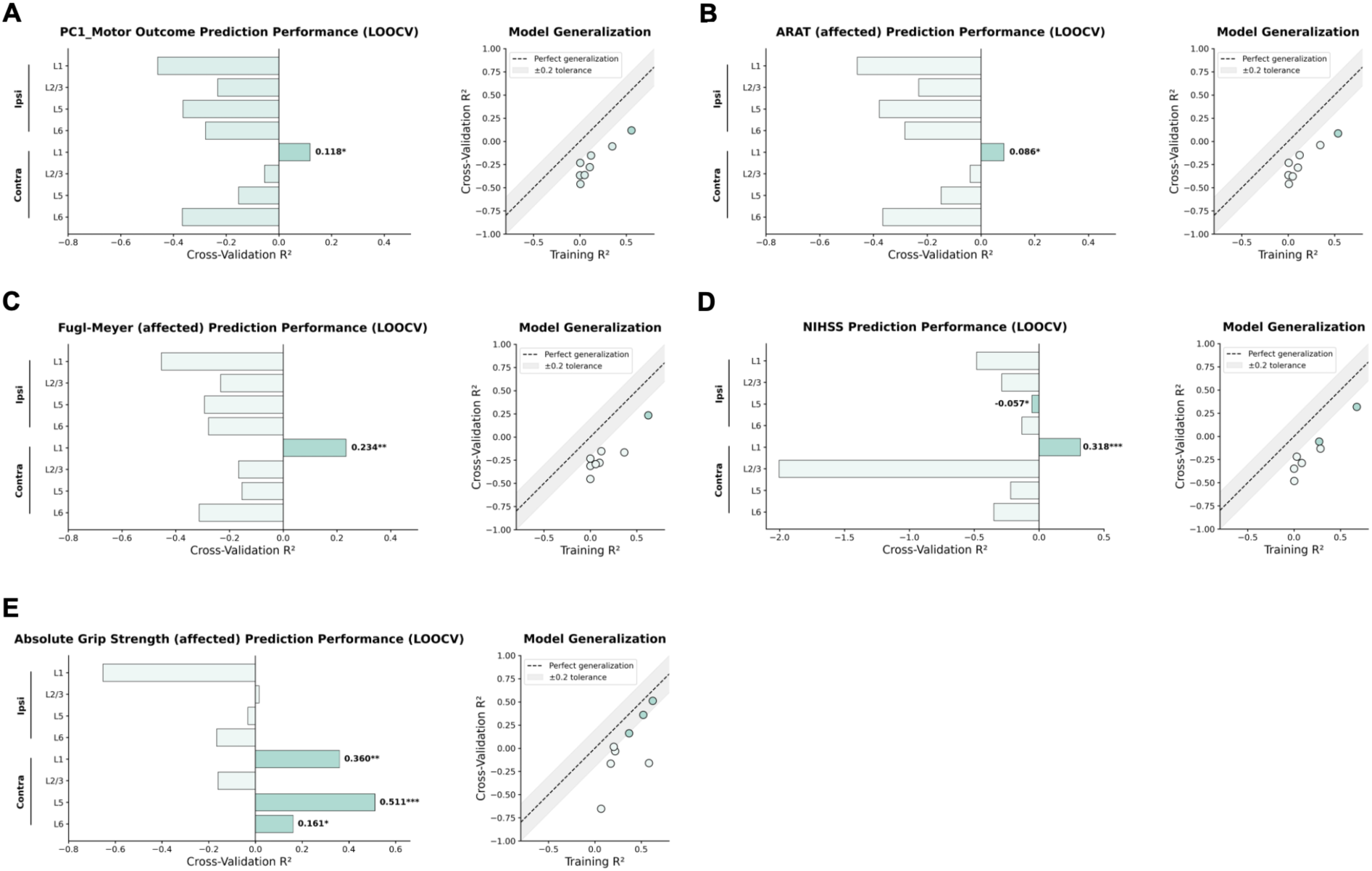
Cross-validation performance for predicting motor outcomes using layer-specific BOLD activation profiles in the lateral hand knob in patients (n = 12). **A**. Leave-one-out cross-validation (LOOCV) performance for predicting PC1 (“motor outcome”) component scores. A linear regression model was fit separately for each cortical layer and hemisphere, using activation as the predictor and PC1 score as the outcome. *Left panel*: Cross-validation R² (1 – SS_residual/SS_total from left-out predictions) for the ipsilesional (Ipsi) and contralesional (Contra) lateral hand knob across cortical layers. *Right panel*: Model generalization comparing training R² versus cross-validation R². Dashed line indicates perfect generalization; shaded area shows ±0.2 tolerance. **B-E**. LOOCV performance (same format as **A**) for predicting individual clinical measures (ARAT and Fugl-Meyer of the affected hand, NIHSS, and absolute grip strength of the affected hand) from layer-specific BOLD activation. \**p* < 0.05; \*\**p* < 0.01; \*\*\**p* < 0.001.

For absolute grip strength of the affected hand, contralesional L1 was also predictive (R² = 0.360, *p* = 0.007; Fig. 3E); however, the strongest cross-validated performance was obtained from the deep output layers, with L5 (R² = 0.511, *p* = 0.0009), and less tense, L6 (R² = 0.161, *p* = 0.036), significantly predicting grip strength. This is consistent with the partial-correlation results, which localized the absolute-grip association to L5 and L6. By contrast, no ipsilesional layer reached significant predictive performance for any measure (all *p* < 0.05, see statistical details in Table S4). Together, these findings again underline the significance of superficial-layer activation in the contralesional lateral hand knob for functional motor outcome after stroke, while indicating that absolute grip strength from the affected hand is additionally predicted by deep output-layer (L5 and L6) activation.

## Discussion

This study provides the first human evidence of layer-specific cortical alterations in chronic stroke using ultra-high-field 7T fMRI. By resolving activation within cortical depth, we identified distinct laminar signatures of stroke-related motor dysfunction in the M1 hand knob that would be invisible to conventional whole-region imaging. The results point to three key findings: (i) ipsilesional impairment affected both input layer (L2/3) and deep output layer (L5) while sparing L1 and L6; (ii) contralesional lateral hand knob showed no group-level overactivation, yet across patients, greater superficial (L1) activation was associated with worse overall motor outcome; and (iii) distinct contralesional compartments related to distinct aspects of motor function, with superficial L1 linked to global motor outcome and deep output layers (L5/L6) linked to grip force.

### Layer-specific dysfunction in the ipsilesional lateral hand knob

Addressing our first aim, stroke-related change in the ipsilesional hand knob was not uniform across depth but was confined to two specific compartments: patients showed significantly reduced activation in L2/3 and L5, while L1 and L6 did not differ from controls. This finding aligns with previous conventional fMRI studies showing decreased ipsilesional motor cortex activation after stroke (Azri et al., 2025; Grefkes & Fink, 2014), but the laminar resolution refines this picture in an informative way, and converges with rodent evidence that these two compartments are selectively affected by focal ischemic injury.

L5 contains the large pyramidal neurons that give rise to corticospinal projections and directly drive voluntary movement (Lemon, 2008; Rathelot & Strick, 2009), so reduced activation likely reflects diminished motor output capacity of the lesioned hemisphere. This is consistent with rodent models of peri-infarct cortex: layer 5 pyramidal neurons show an early loss of dendritic spines that only partially recovers over subsequent weeks (Brown et al., 2008; Mostany et al., 2010), however, their dendritic arbors show no evidence of compensatory growth even when followed through the chronic phase, instead undergoing a two-step pruning process of initial retraction followed by branch loss (Mostany & Portera-Cailliau, 2011). This contrasts with the trajectory of fine-scale spine density, which recovers fully by 2–3 months post-stroke (Mostany et al., 2010), indicating that L5 dendritic arbor architecture, but not spine density per se, remains persistently altered into the chronic phase.

L2/3, by contrast, receives cortico-cortical inputs from premotor, supplementary motor, and somatosensory regions, serving as a site of sensorimotor integration (Mao et al., 2011; McColgan et al., 2020). Its reduced activation may indicate impaired integration of descending motor commands with cortical feedback. This compartment, too, is disrupted early after stroke in rodents, with peri-infarct layer 2/3 pyramidal neurons showing rapid dendritic spine loss within hours of injury (Brown et al., 2008) and elevated tonic GABAergic inhibition emerging within the first week and persisting through at least 21 days post-stroke (Alasoadura et al., 2024; Clarkson et al., 2010).

The finding that both input and output layers are affected suggests that chronic stroke compromises both loops of the microcircuit in M1 rather than a single stage, consistent with models of distributed motor network dysfunction following focal lesions (Campos et al., 2023; Grefkes & Fink, 2014). This is distinct from the pattern reported in focal hand dystonia, where Huber et al. (2023) observed increased superficial layer activity and decreased deep layer activity, interpreted as reduced surround inhibition or excessive sensory feedback with impaired motor output. Rather than an imbalance between input and output processing, stroke reduced both the L2/3 input and L5 output activation, reflecting a more global suppression of motor circuit function. This distinction may reflect the differences in pathophysiology: primary dystonia is a functional network disorder that occurs in the absence of structural lesions (Gill et al., 2023), whereas stroke arises from structural damage and disconnection (Griffis et al., 2019; Salvalaggio et al., 2020).

### Reconsidering the role of the contralesional hand knob

Our second aim was to assess whether the contralesional overactivation reported at 3T would replicate at 7T and, if so, whether it was layer-specific. At the group level, we did not find contralesional overactivation at any layer during affected-hand tapping in patients compared to controls. Instead of a difference in mean activation, the interesting finding was how its activation covaried with behaviour: among patients, greater superficial (L1) activation in the contralesional hand knob was associated with worse motor function. This held across the PCA-derived motor-outcome component (PC1) and, in a convergent fashion, across individual measures: higher contralesional L1 activation was accompanied by lower ARAT and Fugl-Meyer scores of the affected hand and higher NIHSS scores.

The laminar localisation to L1 is mechanistically suggestive. L1 is the primary recipient of long-range cortico-cortical feedback projections and transcallosal inputs, and contains the apical dendritic tufts of pyramidal output neurons located in deeper layers (Ledderose et al., 2023; Schuman et al., 2021). This layer has been proposed as the center for top-down modulation and contextual integration of motor commands. Activity in Layer 1 reflects inputs to—rather than outputs from—the motor cortex, targeting the distal dendrites of Layer 5 corticospinal neurons (M. Larkum, 2013; M. E. Larkum et al., 1999). Thus, excessive L1 activation in the contralesional hemisphere may represent aberrant feedback or interhemispheric signaling that modulates ipsilesional motor output through transcallosal pathways, rather than direct compensatory motor commands. This interpretation aligns with models of maladaptive interhemispheric inhibition, in which contralesional activity interferes with ipsilesional motor reorganisation (Campos et al., 2023; Murase et al., 2004), and with clinical observations that persistent contralesional overactivity in the chronic phase shows poor recovery, and that interfering with contralesional M1 at early stages after stroke can improved motor performance (Grefkes & Fink, 2014; Jones, 2017; Mustin et al., 2024; Takeuchi et al., 2005). Animal models provide mechanistic insight into these observations. Following a cortical stroke, corticospinal tract fibers originating from the contralesional motor cortex sprout across the midline and reinnervate denervated spinal cord regions (Liu et al., 2008). Transcallosal projections from contralesional cortex can reinnervate peri-infarct tissue, but this process may form aberrant connections that hinder rather than support recovery through abnormal interhemispheric inhibition (Murase et al., 2004).

### Distinct contralesional layers track global outcome and grip force

Within the contralesional hand knob, two functionally distinct relationships emerged at different cortical depths. As described above, superficial L1 activation tracked global motor impairment (PC1, ARAT and Fugl-Meyer of the affected hand, NIHSS). Grip force showed a different laminar signature: after accounting for the correlation between absolute and relative grip via partial correlations, the association with elevated contralesional activation was specific to absolute grip strength and localised to the deep output layers (L5/L6), while the apparent L2/3– relative-grip relationship reflected shared variance with absolute strength rather than an independent effect. This produces a layer-specific double dissociation within the contralesional hemisphere: L1 plausibly reflecting maladaptive feedback or interhemispheric signalling (as above), and L5/L6 the engagement of corticospinal output machinery, potentially reflecting greater reliance on contralesional output in patients with weaker affected-hand grip.

This interpretation is consistent with a broader literature implicating the contralesional corticospinal system as a compensatory but incompletely effective route for impaired grip force after stroke. In humans, the coupling between brain activity and peak grip force shifts away from ipsilesional M1 toward bilateral premotor and contralesional dorsal premotor regions specifically in patients with greater corticospinal tract damage, indicating that this contralesional shift is a marker of ipsilesional compromise rather than a route to restored force output (Ward et al., 2007). Anatomical studies support the biological plausibility of such a route. In rodents, contralesional corticospinal fibers sprout into the denervated spinal cord after stroke, and in macaques, contralesional corticospinal tract (CST) axons reorganize to form new connections onto spinal motoneurons during recovery of precision grip (Sawada et al., 2023). Causal silencing of these sprouted, ipsilaterally projecting contralesional fibers disrupts recovered grasping performance in rodents (Wahl et al., 2017), confirming their functional relevance even when recruitment remains an imperfect substitute for the lost ipsilesional pathway. Within this framework, the negative correlation between contralesional L5 activation and absolute grip strength observed here may reflect greater reliance on this compensatory route in patients with more severe corticospinal damage.

### Laminar biomarkers: predictive value, clinical potential, and limitations

The correlational dissociation was mirrored in cross-validated prediction: contralesional L1 activation predicted global motor outcome (PC1, ARAT, Fugl-Meyer of the affected hand, and NIHSS), while absolute grip strength of the affected hand was best predicted by the deep output layers (L5 and, to a lesser extent, L6). No ipsilesional layer reached significance for any measure. This double dissociation reinforces that these compartments carry functionally distinct information rather than a shared “activation” signal.

This distinction has practical implications. Whole-region motor cortex activation collapses these two dissociable processes into a single value, obscuring whether a given patient’s impairment reflects disrupted sensorimotor integration, compromised corticospinal output, or both. A layer-resolved profile could instead flag which mechanism predominates in a given patient. Whether this could eventually inform treatment selection (e.g., prioritizing sensory-feedback-based versus corticospinal-excitability-targeted interventions) remains speculative and would require prospective studies linking laminar signatures to treatment response. Layer-resolved fMRI biomarkers could thus complement, rather than replace, established prognostic markers such as lesion load and corticospinal tract integrity (Boyd et al., 2017; Stinear, 2017).

These results should be interpreted with caution, given several limitations: our small sample size, though sufficient for detecting robust effects, limits generalizability; and our focus on finger tapping tasks in the lateral hand knob may not generalize to other tasks or regions. Despite these constraints, the clear predictive relationships between layer-specific activation and motor outcomes provide initial evidence that warrants validation in larger cohorts. Future longitudinal studies that track laminar changes from the acute to the chronic phase could reveal critical time windows for intervention.

## Conclusion

By resolving motor cortex activation across cortical layers in living stroke patients, this study shows that chronic stroke produces alterations that are organized by lamina: a dual input–output reduction (L2/3 and L5) within the ipsilesional hand knob, and a functionally graded contralesional signature in which superficial feedback-layer activation tracks global impairment and deep output-layer activation tracks grip force. These findings advance the understanding of post-stroke reorganisation beyond the whole-region level and indicate that laminar fMRI may serve as a tool for characterising sensorimotor circuit dysfunction after stroke. Although current non-invasive neuromodulation cannot target individual layers, the markers identified here could support prognosis, patient stratification, and monitoring of response to circuit-level interventions.

## Data Availability

All data produced in the present study are available upon reasonable request to the authors

## Acknowledgments

This work was funded by the Deutsche Forschungsgemeinschaft (DFG, German Research Foundation) – Project-ID 431549029 – SFB 1451 (project C05).

