## Supplementary material for "Layer-specific cortical signatures uncover a sensory origin of post-stroke motor dysfunction": Table S1-S4

**Table S1**. Statistical results of the comparison of layer-specific activation between control and patient groups in the lateral hand knob during finger tapping of the affected/right hand.

| Hemisphere | Layer | U | p_perm | p_FDR | Cohen's d |
| --- | --- | --- | --- | --- | --- |
| Ipsilesional (left) | L1 | 111 | 0.088 | 0.118 | 0.42 |
|  | L2/3 | 125 | 0.018 | 0.043 | 0.63 |
|  | L5 | 124 | 0.021 | 0.043 | 0.70 |
|  | L6 | 93 | 0.330 | 0.330 | 0.25 |
| Contralesional (right) | L1 | 113 | 0.071 | 0.286 | 0.52 |
|  | L2/3 | 104 | 0.163 | 0.327 | 0.02 |
|  | L5 | 79 | 0.601 | 0.653 | -0.16 |
|  | L6 | 76 | 0.653 | 0.653 | -0.12 |

(p_perm = permutation p-value, 5000 iterations; p_FDR = p-value after FDR correction across the four layers within each hemisphere)

**Table S2**. Statistical results of Pearson correlation between PC1 and PC2 component scores and layer-specific activation during finger tapping of the affected hand in the ipsilesional and contralesional lateral hand knob in the patient group.

| Components | Hemisphere | Layer | r | p | p (FDR) |
| --- | --- | --- | --- | --- | --- |
| PC1 | Ipsilesional | L1 | 0.078 | 0.820 | 0.853 |
|  |  | L2/3 | 0.063 | 0.853 | 0.853 |
|  |  | L5 | -0.226 | 0.504 | 0.853 |
|  |  | L6 | -0.327 | 0.326 | 0.853 |
|  | Contralesional | L1 | -0.746 | 0.005 | 0.021 |
|  |  | L2/3 | -0.591 | 0.042 | 0.086 |
|  |  | L5 | -0.347 | 0.269 | 0.359 |
|  |  | L6 | -0.038 | 0.908 | 0.908 |
| PC2 | Ipsilesional | L1 | -0.543 | 0.084 | 0.169 |
|  |  | L2/3 | -0.107 | 0.755 | 0.755 |
|  |  | L5 | 0.456 | 0.159 | 0.212 |
|  |  | L6 | 0.552 | 0.079 | 0.169 |
|  | Contralesional | L1 | 0.039 | 0.904 | 0.922 |
|  |  | L2/3 | -0.081 | 0.803 | 0.922 |
|  |  | L5 | 0.032 | 0.922 | 0.922 |
|  |  | L6 | 0.180 | 0.575 | 0.922 |

(r = Pearson correlation coefficient; p = p-value; p_FDR = p-value after FDR correction across the four layers within each hemisphere)

**Table S3**. Statistical results of Pearson correlation between individual clinical measures and layer-specific activation during finger tapping of the affected hand in the lateral hand knob in the patient group.

| Measure | Hemisphere | Layer | r | p | p_FDR |
| --- | --- | --- | --- | --- | --- |
| ARAT (affected) | Ipsilesional | L1 | 0.087 | 0.798 | 0.829 |
|  |  | L2/3 | 0.074 | 0.829 | 0.829 |
|  |  | L5 | −0.220 | 0.515 | 0.829 |
|  |  | L6 | −0.323 | 0.332 | 0.829 |
|  | Contralesional | L1 | −0.733 | 0.007 | 0.027 |
|  |  | L2/3 | −0.588 | 0.044 | 0.089 |
|  |  | L5 | −0.351 | 0.263 | 0.351 |
|  |  | L6 | −0.049 | 0.879 | 0.879 |
| Fugl-Meyer (affected) | Ipsilesional | L1 | −0.006 | 0.987 | 0.987 |
|  |  | L2/3 | −0.015 | 0.966 | 0.987 |
|  |  | L5 | −0.240 | 0.476 | 0.953 |
|  |  | L6 | −0.315 | 0.346 | 0.953 |
|  | Contralesional | L1 | −0.790 | 0.002 | 0.009 |
|  |  | L2/3 | −0.605 | 0.037 | 0.074 |
|  |  | L5 | −0.344 | 0.273 | 0.364 |
|  |  | L6 | −0.019 | 0.953 | 0.953 |
| NIHSS | Ipsilesional | L1 | −0.046 | 0.892 | 0.892 |
|  |  | L2/3 | 0.289 | 0.388 | 0.517 |
|  |  | L5 | 0.521 | 0.101 | 0.201 |
|  |  | L6 | 0.534 | 0.091 | 0.201 |
|  | Contralesional | L1 | 0.823 | 0.001 | 0.004 |
|  |  | L2/3 | 0.309 | 0.329 | 0.659 |
|  |  | L5 | 0.173 | 0.592 | 0.789 |
|  |  | L6 | 0.017 | 0.959 | 0.959 |
| Absolute grip strength (affected) | Ipsilesional | L1 | −0.260 | 0.440 | 0.440 |
|  |  | L2/3 | −0.452 | 0.163 | 0.275 |
|  |  | L5 | −0.468 | 0.147 | 0.275 |
|  |  | L6 | −0.414 | 0.206 | 0.275 |
|  | Contralesional | L1 | −0.723 | 0.008 | 0.011 |
|  |  | L2/3 | −0.764 | 0.004 | 0.008 |
|  |  | L5 | −0.790 | 0.002 | 0.008 |
|  |  | L6 | −0.608 | 0.036 | 0.036 |
| Relative grip strength | Ipsilesional | L1 | −0.313 | 0.348 | 0.618 |
|  |  | L2/3 | −0.140 | 0.682 | 0.682 |
|  |  | L5 | −0.247 | 0.464 | 0.618 |
|  |  | L6 | −0.316 | 0.344 | 0.618 |
|  | Contralesional | L1 | −0.626 | 0.029 | 0.059 |
|  |  | L2/3 | −0.704 | 0.011 | 0.043 |
|  |  | L5 | −0.383 | 0.219 | 0.292 |
|  |  | L6 | −0.048 | 0.882 | 0.882 |

(r = Pearson correlation coefficient; p = p-value; p_FDR = p-value after FDR correction across the four layers within each hemisphere)

**Table S4**. LOOCV prediction results (univariate models predicting each outcome from single-layer activation in the ipsilesional/contralesional lateral hand knob during finger tapping of the affected hand).

| Outcome | Hemisphere | Layer | Training R² | CV R² | Overfit gap | p_perm |
| --- | --- | --- | --- | --- | --- | --- |
| PC1 (motor outcome) | Ipsilesional | L1 | 0.006 | −0.460 | 0.466 | 0.746 |
|  |  | L2/3 | 0.004 | −0.232 | 0.236 | 0.124 |
|  |  | L5 | 0.051 | −0.364 | 0.415 | 0.423 |
|  |  | L6 | 0.107 | −0.278 | 0.385 | 0.219 |
|  | Contralesional | L1 | 0.557 | 0.118 | 0.438 | 0.030 |
|  |  | L2/3 | 0.350 | −0.054 | 0.404 | 0.045 |
|  |  | L5 | 0.120 | −0.153 | 0.273 | 0.104 |
|  |  | L6 | 0.001 | −0.367 | 0.368 | 0.587 |
| ARAT (affected) | Ipsilesional | L1 | 0.008 | −0.460 | 0.468 | 0.735 |
|  |  | L2/3 | 0.005 | −0.232 | 0.237 | 0.153 |
|  |  | L5 | 0.049 | −0.378 | 0.427 | 0.466 |
|  |  | L6 | 0.105 | −0.283 | 0.388 | 0.267 |
|  | Contralesional | L1 | 0.537 | 0.086 | 0.452 | 0.016 |
|  |  | L2/3 | 0.346 | −0.040 | 0.386 | 0.050 |
|  |  | L5 | 0.123 | −0.149 | 0.272 | 0.098 |
|  |  | L6 | 0.002 | −0.365 | 0.367 | 0.614 |
| Fugl-Meyer (affected) | Ipsilesional | L1 | 0.000 | −0.453 | 0.453 | 0.730 |
|  |  | L2/3 | 0.000 | −0.233 | 0.233 | 0.144 |
|  |  | L5 | 0.058 | −0.293 | 0.351 | 0.262 |
|  |  | L6 | 0.099 | −0.278 | 0.377 | 0.234 |
|  | Contralesional | L1 | 0.624 | 0.234 | 0.391 | 0.006 |
|  |  | L2/3 | 0.366 | −0.166 | 0.533 | 0.067 |
|  |  | L5 | 0.118 | −0.154 | 0.272 | 0.075 |
|  |  | L6 | 0.000 | −0.312 | 0.313 | 0.520 |
| NIHSS | Ipsilesional | L1 | 0.002 | −0.482 | 0.484 | 0.755 |
|  |  | L2/3 | 0.084 | −0.288 | 0.372 | 0.290 |
|  |  | L5 | 0.271 | −0.057 | 0.328 | 0.021 |
|  |  | L6 | 0.285 | −0.133 | 0.418 | 0.067 |
|  | Contralesional | L1 | 0.677 | 0.318 | 0.359 | 0.001 |
|  |  | L2/3 | 0.095 | −2.003 | 2.098 | 0.984 |
|  |  | L5 | 0.030 | −0.219 | 0.249 | 0.140 |
|  |  | L6 | 0.000 | −0.349 | 0.349 | 0.601 |
| Absolute grip strength (affected) | Ipsilesional | L1 | 0.068 | −0.653 | 0.721 | 0.955 |
|  |  | L2/3 | 0.204 | 0.015 | 0.189 | 0.091 |
|  |  | L5 | 0.219 | −0.032 | 0.251 | 0.103 |
|  |  | L6 | 0.171 | −0.166 | 0.337 | 0.179 |
|  | Contralesional | L1 | 0.522 | 0.360 | 0.162 | 0.007 |
|  |  | L2/3 | 0.583 | −0.160 | 0.744 | 0.184 |
|  |  | L5 | 0.624 | 0.511 | 0.113 | 0.001 |
|  |  | L6 | 0.369 | 0.161 | 0.208 | 0.036 |

(R²_CV = cross-validated R²; p_perm = permutation p-value, 10,000 iterations)
